# Serial infection with SARS-CoV-2 Omicron BA.1 and BA.2 following three-dose COVID-19 vaccination

**DOI:** 10.1101/2022.05.19.22275026

**Authors:** Hope R. Lapointe, Francis Mwimanzi, Peter K. Cheung, Yurou Sang, Fatima Yaseen, Rebecca Kalikawe, Sneha Datwani, Rachel Waterworth, Gisele Umviligihozo, Siobhan Ennis, Landon Young, Winnie Dong, Don Kirkby, Laura Burns, Victor Leung, Daniel T. Holmes, Mari L. DeMarco, Janet Simons, Nancy Matic, Julio S.G. Montaner, Chanson J. Brumme, Natalie Prystajecky, Masahiro Niikura, Christopher F. Lowe, Marc G. Romney, Mark A. Brockman, Zabrina L. Brumme

**Author notes:** denote equal contribution (Hope R. Lapointe, Francis Mwimanzi, Marc G. Romney, Mark A. Brockman, Zabrina L. Brumme). **Corresponding author:** Zabrina L. Brumme, Ph.D., Professor, Faculty of Health Sciences, Simon Fraser University, 8888 University Drive, Burnaby, BC, Canada, V5A 1S6.

## Abstract

SARS-CoV-2 Omicron infections are common among individuals who are vaccinated or have recovered from prior variant infection, but few reports have documented serial Omicron infections. We characterized SARS-CoV-2 humoral responses in a healthy young person who acquired laboratory-confirmed Omicron BA.1.15 ten weeks after a third dose of BNT162b2, and BA.2 thirteen weeks later. Responses were compared to those of 124 COVID-19 naive vaccinees. One month after the second and third vaccine doses, the participant’s wild-type and BA.1-specific IgG, ACE2 competition and virus neutralization activities were average for a COVID-19 naive triple-vaccinated individual. BA.1 infection boosted the participant’s responses to the cohort ≥95th percentile, but even this strong “hybrid” immunity failed to protect against BA.2. Moreover, reinfection increased BA.1 and BA.2-specific responses only modestly. Results illustrate the risk of Omicron infection in fully vaccinated individuals and highlight the importance of personal and public health measures as vaccine-induced immune responses wane.

## INTRODUCTION

SARS-CoV-2 infections, predominantly fueled by the Omicron (B.1.1.529) variant, are increasingly common among individuals who are vaccinated and/or have recovered from prior infections (1-3). Globally, the highly transmissible and immune evasive Omicron variant has rapidly overtaken the previously dominant Delta variant (3-7), and the original Omicron BA.1 strain is being outcompeted by newer Omicron sub-lineages BA.2, BA.3, BA.4 and BA.5 (8, 9). In British Columbia (BC), Canada, Omicron BA.1 had overtaken Delta by December 2021 and BA.2 had largely outcompeted BA.1 by March 2022 (10, 11).

COVID-19 vaccine coverage in BC is relatively high, with 93%, 90% and 57% of individuals aged 12 years or older having received one, two and three COVID-19 immunizations, respectively (12). Persons at elevated risk of severe COVID-19 are also eligible for fourth doses (13). Despite this, the province experienced fifth and sixth waves of COVID-19, dominated by BA.1 and BA.2, respectively, as public health measures were gradually relaxed (10, 11). Indeed, it is estimated that between December 2021 and March 2022, nearly half of British Columbians experienced a SARS-CoV-2 infection, likely due to Omicron (14, 15).

While several reports have examined post-vaccination Omicron infections, or Omicron reinfections following exposure to prior variants (16-23), we are aware of only one study that assessed repeat Omicron infection incidence through viral genomic surveillance (24). No studies appear to have investigated the vaccine- and infection-induced immune responses after serial Omicron infections. Here, we longitudinally characterize SARS-CoV-2 humoral responses in a healthy young person who experienced serial BA.1 and BA.2 Omicron infections following three-dose COVID-19 mRNA vaccination. Responses were compared to those of 124 COVID-19-naive vaccinees over the same period. Taken together with existing literature, our results suggest that vaccination provides limited protection against infection and/or reinfection by Omicron variants.

## METHODS

### Observational COVID-19 vaccine cohort and SARS-CoV-2 infection monitoring

In December 2020, we established a prospective longitudinal study in Vancouver, Canada, to examine SARS-CoV-2 specific humoral immune responses following vaccination with BNT162b2 (Comirnaty; BioNTech/Pfizer) or mRNA-1273 (Spikevax; Moderna) in a cohort of 151 adults aged 24-98 years (described in (25, 26)). Serum and plasma were collected longitudinally up to 6 months following the third dose (Figure 1A). At each visit, serum was tested for the presence of SARS-CoV-2 anti-nucleocapsid (N) antibodies, which indicate seroconversion following infection, using the Elecsys Anti-SARS-CoV-2 assay on a Cobas e601 module analyzer (Roche Diagnostics). In addition to the case participant, immune measures from a comparison group of 124 additional cohort participants who remained anti-N seronegative up until at least one month post-third vaccine dose are included for context.

**Figure 1.**
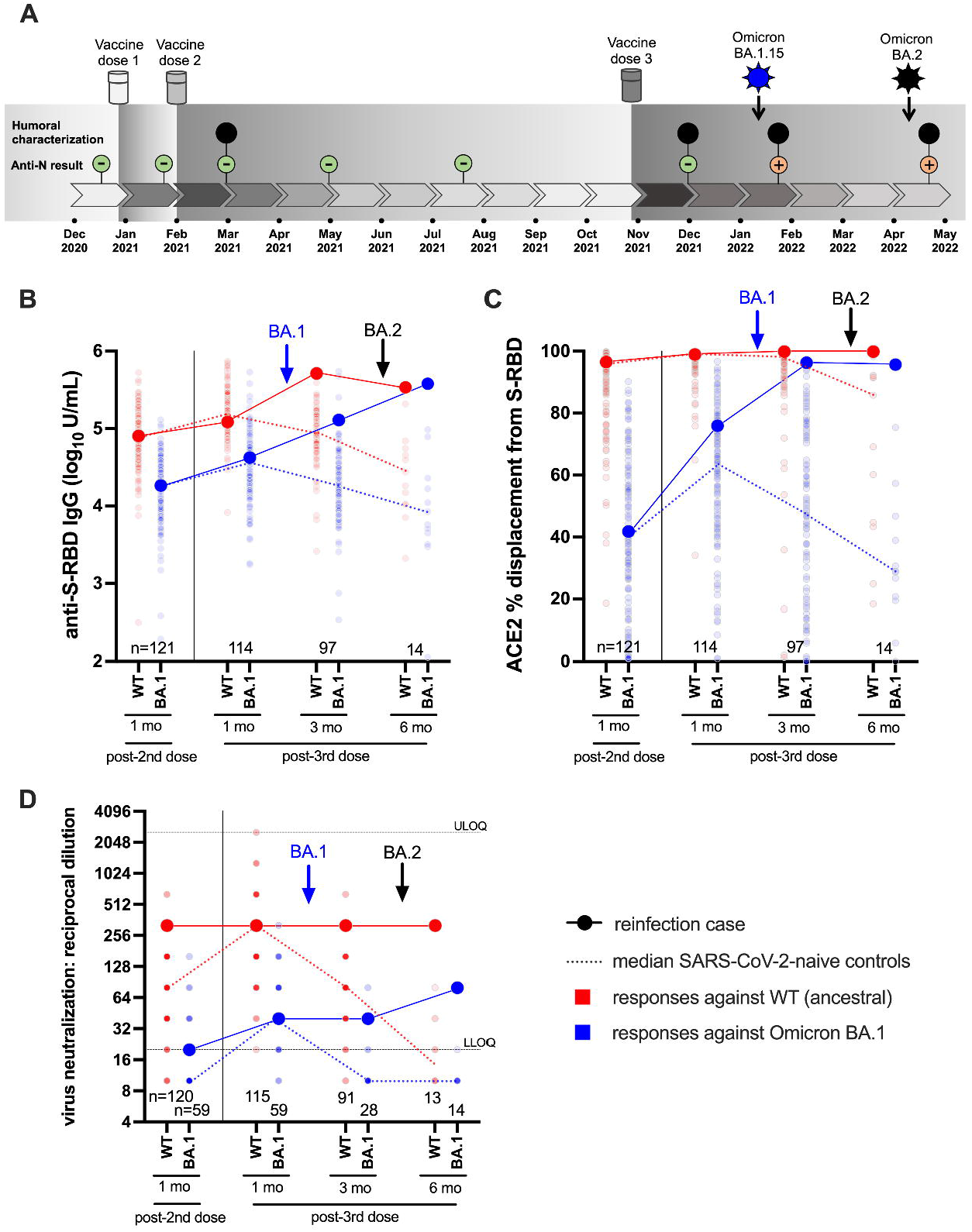
Case participant history and longitudinal humoral responses against wild-type and Omicron BA.1 SARS-CoV-2. Panel A: Case participant timeline. Immunizations and SARS-CoV-2 Omicron infection history are shown at the top. Longitudinal SARS-CoV-2 anti-N serology results are shown in small green (anti-N negative) or orange (anti-N positive) circles. Large black circles denote time points where additional humoral functions, shown in panels below, were measured. Panel B: Longitudinal anti-S-RBD IgG log10 concentrations in case participant (large circles) versus the comparison group of SARS-CoV-2-naive individuals (small circles) at various time points following two- and three-dose COVID-19 vaccination. Wild-type (WT) specific anti-S-RBD responses are shown in red; Omicron BA.1-specific ones are shown in blue. Matching solid lines connect the participant’s longitudinal values, while dotted lines connect the median values for the comparison group. Approximate times of BA.1 and BA.2 infections are shown with arrows. Total Ns (including the case participant) are shown at the bottom of the plot. Later time points have smaller Ns because some control participants were censored due to post-vaccination SARS-CoV-2 infection or had not yet completed the visit. Panel C: same as previous, but for longitudinal ACE2% displacement function from wild-type (red) and BA.1 (blue) S-RBDs. Panel D: same as previous, but for longitudinal live virus neutralization function against wild-type (red) and BA.1 (blue) strains. ULOQ/LLOQ: upper/lower limit of quantification.

### Ethics approval

All participants provided written informed consent. This study was approved by the University of British Columbia/Providence Health Care and Simon Fraser University Research Ethics Boards (protocol H20-03906).

### SARS-CoV-2 diagnostics and lineage confirmation

Diagnostic samples from the case participant’s two SARS-CoV-2 infections were tested at the St. Paul’s Hospital Virology Laboratory using the cobas® SARS-CoV-2 Test which targets conserved regions within the Orf1a/b and E genes (Roche Diagnostics) followed by screening using a real-time reverse transcription (RT)-PCR based algorithm for SARS-CoV-2 lineage classification that is frequently updated to detect emerging variants (27, 28). Following this, the diagnostic samples were subjected to full-genome SARS-CoV-2 sequencing in two independent laboratories: the BC Centre for Disease Control, the provincial laboratory that performs all SARS-CoV-2 sequencing for epidemiological surveillance, and the BC Centre for Excellence in HIV/AIDS. Both laboratories use the Illumina platform. The SARS-CoV-2 full genome sequences for the participant’s BA.1.15 and subsequent BA.2 infections have been submitted to GISAID (Accession ID EPI_ISL_12767799 and EPI_ISL_12662303, respectively).

### Binding antibody assays

We quantified anti-Spike Receptor Binding Domain (RBD) binding IgG concentrations in serum using the V-plex SARS-CoV-2 (IgG) Panel 22 ELISA kit (Meso Scale Diagnostics), which features wild-type and Omicron BA.1 RBD antigens. For a subset of participants, Anti-Spike binding IgG concentrations in serum were also quantified using the V-plex SARS-CoV-2 (IgG) Panel 25 ELISA kit (Meso Scale Diagnostics), which features full-length S antigens from wild-type, Omicron BA.1 and Omicron BA.2. At the time of analysis, no panel featuring Omicron BA.2 RBD was offered by the manufacturer. Both assays were performed on a Meso QuickPlex SQ120 instrument, with sera diluted 1:10000. Results are reported in arbitrary Units/mL.

### ACE2 competition assays

We assessed the ability of serum antibodies to block the wild-type and Omicron BA.1 RBD-ACE2 receptor interaction by competition ELISA (Panel 22 V-plex SARS-CoV-2 [ACE2]; Meso Scale Diagnostics). For a subset of participants, we also assessed the ability of serum antibodies to block the wild-type, BA.1 and BA.2 Spike-ACE2 receptor interaction using the same methods (Panel 25 V-plex SARS-CoV-2 [ACE2]). Both assays were performed on a Meso QuickPlex SQ120 instrument, with sera diluted 1:40. Results are reported as % ACE2 displacement.

### Live virus neutralization assays

Neutralizing activity in plasma was examined in live SARS-CoV-2 assays using a wild-type isolate (USA-WA1/2020; BEI Resources) and a local Omicron BA.1 isolate (GISAID Accession # EPI_ISL_9805779) on VeroE6-TMPRSS2 (JCRB-1819) target cells. Viral stock was adjusted to 50 TCID50/200 µl in Dulbecco’s Modified Eagle Medium in the presence of serial 2-fold plasma dilutions (from 1/20 to 1/2560), incubated at 4°C for 1 hour and added to target cells in 96-well plates in triplicate. Cultures were maintained at 37°C with 5% CO2 and the appearance of viral cytopathic effect (CPE) was recorded three days post-infection. Neutralizing activity is reported as the reciprocal of the highest plasma dilution able to prevent CPE in all triplicate wells. Samples exhibiting partial or no neutralization at 1/20 dilution were defined as below the limit of quantification (BLOQ).

### Statistical analysis

Data visualization and statistical analysis was conducted in Prism v9.2.0 (GraphPad).

## RESULTS

### Case participant SARS-CoV-2 vaccination and infection timeline

The participant was a frontline health care worker in their early 30s with no chronic health conditions. They received three doses of mRNA vaccine (all BNT162b2; 30mcg) in late December 2020, early February 2021 and late October 2021 (Figure 1A). All blood samples collected up to one month following the third immunization were anti-N seronegative.

In early January 2022, ten weeks after the third immunization, the participant experienced moderate symptoms including sore throat, fatigue, congestion, body aches, severe headaches, loss of taste and smell, coughing, shortness of breath and nausea. Symptoms, primarily cough, intensified in the second week after diagnosis requiring corticosteroid therapy. By the third week, symptoms had subsided except for shortness of breath and fatigue, with minimal improvement from short- and long-acting bronchodilating agents. A saline gargle collected in early January 2022 tested positive on the cobas® SARS-CoV-2 Test with a cycle threshold (Ct) value of 21 for both Orf1a/b and E gene targets. Real-time RT-PCR-based molecular screening identified the infection as Omicron BA.1, with subsequent full-genome viral sequencing confirming the specific lineage as BA.1.15.

In early April 2022, 13 weeks following the BA.1 infection (and 23 weeks following the third immunization) the participant experienced a different, more mild symptom profile compared to their first infection, consisting of a sore throat, fever, body aches, headaches, and diarrhea. No change in sense of taste or smell was noted. The participant noted persisting weakness, fatigue and mental fog, as well as severe long-term, treatment-resistant shortness of breath triggered by mild activities or exercises. A nasopharyngeal swab collected in early April 2022 tested positive on the cobas® SARS-CoV-2 Test with Ct values of 24 (Orf1a/b) and 23 (E). This second infection was identified as BA.2 by molecular screening and confirmed by full-genome viral sequencing.

### Longitudinal humoral responses to wild-type and Omicron BA.1 variants

We began by investigating the magnitude of the participant’s humoral immune responses following immunization, in context of a control group of 124 COVID-naïve individuals who were vaccinated during the same period. The comparison group was 74% female with a median age of 57 (Interquartile Range [IQR] 38-76) years. We quantified antibody responses to wild-type and Omicron BA.1 strains in the participant and the comparison group at one month after the second and third vaccine doses, as these time points should capture peak responses post-vaccination (Figures 1B-D).

One month post-second dose, the participant’s wild-type and BA.1-specific RBD IgG concentrations were 4.90 and 4.26 log_10_ U/mL respectively, which were equivalent to the 53^rd^ and 50^th^ percentile values of the comparator cohort (Figure 1B). One month post-third dose, their wild-type and BA.1-specific RBD IgG concentrations had increased to 5.08 and 4.63 log_10_ U/mL respectively, equivalent to the 36^th^ and 60^th^ percentile values of the cohort. Similarly, one month post-second dose, their ability to disrupt the interaction between the ACE2 receptor and the wild-type and BA.1 RBDs were 97% (57^th^ percentile) and 42% (55^th^ percentile) respectively (Figure 1C). At one month post-third dose, their wild-type- and BA.1-specific RBD-ACE2 displacement activities had increased to 99% (74^th^ percentile) and 76% (73^rd^ percentile) respectively. Finally, at one month post-second dose, the participant’s plasma neutralized wild-type and BA.1 SARS-CoV-2 at reciprocal dilutions of 320 and 20, which were equivalent to the 97^th^ and 76^th^ percentile values of the cohort. At one month post-third dose, their wild-type and BA.1 neutralization titers were 320 (78^th^ percentile) and 40 (59^th^ percentile) respectively. These results indicate that the participant’s overall vaccine responses were typical of the cohort, but nevertheless insufficient to prevent infection by BA.1 approximately six weeks later.

Seventeen days after testing positive with BA.1 (which coincided with a three-month post-third-dose study visit), the participant’s wild-type and BA.1-specific responses were boosted substantially, reaching the cohort 95th percentile for most measures at this time when immune responses had begun to decline in the comparator cohort (Figures 1B-1D). Their wild-type RBD IgG concentration increased to 5.71 log_10_ U/mL, while their BA.1-specific RBD IgG concentration increased to 5.11 log_10_ U/mL (Figure 1B). For context, these values would have placed the participant in the 95th and 93rd percentile of “peak” cohort values, measured at one month post-third vaccine dose. Similarly, their wild-type-specific RBD-ACE2 competition activity remained high at 99.9%, while their BA.1-specific RBD-ACE2 competition activity increased to 96.3%. For context, these values were equivalent to the 99th percentiles of peak cohort values one month after three vaccine doses (Figure 1C). Their wild-type and BA.1-specific neutralization values held at 320 and 40, respectively, equivalent to the 78^th^ and 59^th^ percentiles of peak cohort values (Figure 1D). These results indicate that BA.1 infection substantially boosted the participant’s humoral response, however this was insufficient to prevent reinfection by BA.2 approximately 10 weeks later.

Sixteen days after testing positive with BA.2 (which coincided with a six-month post-third-dose study visit), the participant’s wild-type-specific responses remained steady or declined slightly (*e*.*g*. RBD IgG) from prior measurements. Nevertheless, most values remained at the cohort 100th percentile at this time point, which is unsurprising given that vaccine-induced responses had declined substantially over this time in the COVID-19 naive comparison group.

By contrast, the BA.2 reinfection had mixed effects on BA.1 responses. While the BA.1-specific RBD IgG concentration rose substantially to 5.58 log_10_ U/mL (whereas the cohort median at this time point was nearly 2 log_10_ lower), no change was seen for BA.1-specific RBD-ACE2 competition, and BA.1 neutralization increased only modestly. The more pronounced impact of BA.2 reinfection was to extend the duration of BA.1-specific responses in the participant, who maintained an RBD-ACE2 competition activity of 95.7% (cohort median 29% at this time point) and a neutralization activity of 80 (cohort median BLOQ at this time point). Nevertheless, despite BA.1 infection and BA.2 reinfection, the participant’s virus neutralization activity against BA.1 at this time point, which represented the highest activity measured during the study, remained 4-fold lower compared to that against the wild-type strain one month following their third vaccine dose (Figure 1D), suggesting that the participant may remain at risk of additional Omicron infection.

### Longitudinal humoral responses to Omicron BA.2

We next characterized BA.2-specific Spike IgG and ACE2 competition activities in a subset of participants (79% Female, median age 59 years) beginning one month following the third vaccine dose (Figure 2). As these analyses focus on whole Spike (rather than RBD antigen), the corresponding wild-type and BA.1-specific responses are also shown for context (Figure 2). We additionally confirmed the (strong) correlations between wild-type- and BA.1-specific RBD and Spike responses in these individuals (all p<0.0001; Figure S1). At one month post-third vaccine dose, the participant displayed wild-type, BA.1 and BA.2-specific Spike IgG concentrations of 5.81, 5.18 and 5.46 log_10_ U/mL respectively (Figure 2A), and ACE2 competition activities of 99.4%, 50.6% and 64.3% respectively (Figure 2B). Their wild-type and BA.2-specific Spike IgG concentrations were broadly average (54^th^ and 68^th^ percentiles respectively), as were their and Spike-ACE2 competition activities (46^th^ and 54^th^ percentiles respectively), though their values for BA.1-specific IgG and BA.1 Spike-ACE2 competition were slightly lower than average (37^th^ and 39^th^ percentiles respectively).

**Figure 2.**
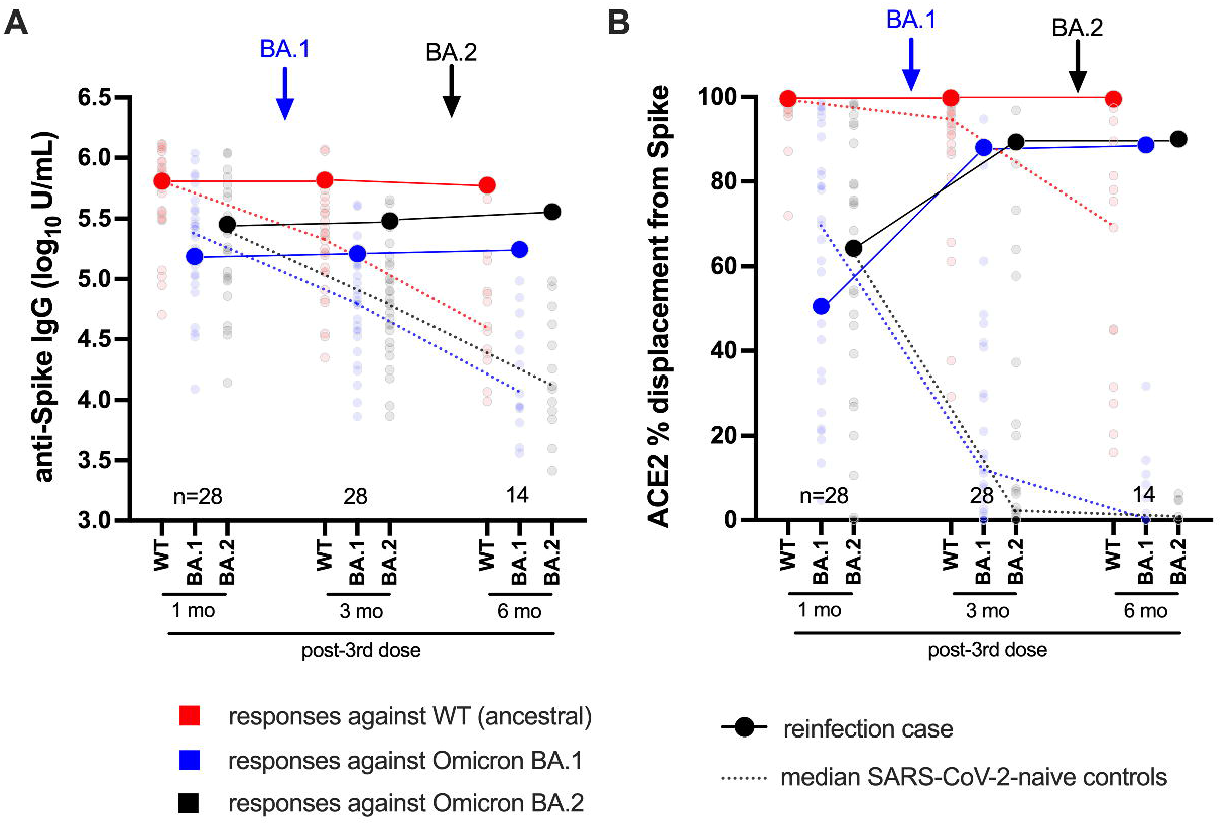
Longitudinal humoral responses against wild-type, BA.1 and BA.2 Spike antigens. Panel A: Anti-Spike IgG log10 concentrations in case participant (large circles) versus the subset of the comparison group of SARS-CoV-2-naive individuals (small circles) at one, three and six months following three-dose COVID-19 vaccination. Wild-type-specific (WT) anti-Spike responses are in red; BA.1-specific ones are in blue; BA.2-specific ones are in black. Matching solid lines connect the participant’s longitudinal values; dotted lines connect the median values for the comparison group. Approximate times of BA.1 and BA.2 infections are shown with arrows. Total Ns (including the case participant) are shown at the bottom of the plot; the final time point has a smaller N because some control participants were censored due to post-vaccination SARS-CoV-2 infection or had not yet completed the visit. Panel B: same as previous, but for longitudinal ACE2% displacement function from wild-type (red), BA.1 (blue) and BA.2 (black) Spike protein.

Following BA.1 infection, the participant’s wild-type, BA.1 and BA.2-specific Spike IgG concentrations increased modestly, to 5.82, 5.22 and 5.48 log10 U/mL respectively (Figure 2A). Though the magnitude of these increases was not as pronounced as those observed for their wild-type- and BA.1-specific RBD IgG concentrations (shown in Figure 1B), these values nevertheless placed them at or above the cohort 85^th^ percentile at this time point, when immune responses had begun to decline in the broader cohort. For context, these values would place the participant in the 57^th^, 39^th^, and 68^th^ percentile of peak cohort values measured at one month post-third vaccine dose. Similar to the ACE2 competition activities measured using RBD antigens (shown in Figure 1C), the participant’s wild-type Spike-ACE2 competition activities remained high at 99.7%, while BA.1 and BA.2 Spike-ACE2 activities rose substantially to 87.9% and 89.3% respectively (Figure 2B); values that represented the 71^st^, 75^th^, and 79^th^ percentiles of peak cohort values at one month post-third vaccine dose.

Following BA.2 infection, the participant’s wild-type Spike IgG concentration declined slightly to 5.77 log_10_ U/mL, whereas their BA.1 and BA.2-specific values increased slightly to 5.25 and 5.56 log_10_ U/mL respectively (Figure 2A). These trends were consistent with their wild-type- and BA.1-specific RBD IgG concentrations (shown in Figure 1B), though of a smaller magnitude. Similar to the ACE2 competition activities measured using RBD antigens (shown in Figure 1C), the participant’s wild-type Spike-ACE2 competition activity remained high (99.5%) after BA.2 infection. BA.1 and BA.2 Spike-ACE2 activities increased, though only marginally, to 88.6% and 90% respectively (Figure 2B).

Together, these results confirm that the participant’s humoral responses to wild-type and Omicron variants were broadly average one month post-third vaccine dose. Moreover, while BA.1 infection boosted Omicron-specific immune responses (highlighted by an increase in BA.1 and BA.2 Spike-ACE2 competition activities), BA.2 reinfection did not substantially augment these activities further but rather extended the duration of these responses.

## DISCUSSION

This study provides detailed humoral characterization in a laboratory-confirmed case of serial infection by SARS-CoV-2 Omicron subvariants BA.1 and BA.2 in an otherwise healthy adult who had received three doses of COVID-19 mRNA vaccine. While data on repeat Omicron infections remain very limited, a recent study from Denmark identified 47 cases of BA.2 reinfection that occurred between 20 and 60 days following BA.1 infection (24). The authors concluded that such events were rare (<0.1% of cases during the brief window of analysis) and more likely to occur among unvaccinated individuals, but further evaluation of the data indicates a majority of reinfection cases were due to BA.2 following BA.1. Moreover, given that the present case participant was one of only 151 enrolees of an observational COVID-19 vaccine study (25, 26), our results suggest that the risk of serial infection with Omicron subvariants may be greater than current assumptions based on low reinfection rates. We note however that the participant’s status as a frontline healthcare worker may have resulted in an increased risk of exposure and (re-)infection over the general population.

Acknowledging that our ability to generalize from a single case is limited, we note that initial vaccine-induced IgG, ACE2 competition and virus neutralization response magnitudes against wild-type and Omicron BA.1 in the participant were comparable to the median values observed in diverse controls who were vaccinated along the same timeline. The observation that average humoral responses to three-dose vaccination failed to protect against Omicron BA.1 infection is consistent with the extremely high rates of community transmission observed in many regions during the recent Omicron-driven pandemic waves. Given that third doses substantially boost humoral responses in individuals of all ages (29-32), the risk of Omicron infection is likely to be higher among individuals who have received fewer than three doses (18, 33) and is likely to increase with time following vaccination due to natural declines in antibody responses (26, 34-37). Additional studies are needed to assess these factors.

While it is perhaps unsurprising that COVID-19 vaccines based on ancestral SARS-CoV-2 sequences will not generate sterilizing immunity against Omicron strains that have evolved to evade host immune responses (4, 38-42), various lines of evidence suggest that “hybrid” immunity resulting from vaccination plus infection provides greater protection against SARS-CoV-2 variants (5, 43), due in part to maturation of Spike-specific antibodies (44-46) and expansion of antiviral T cells (47-52). In light of this, we note that symptomatic BA.1 infection boosted vaccine-induced humoral responses against both BA.1 and BA.2 in our case participant, but the heightened response nevertheless failed to prevent subsequent symptomatic infection by BA.2. Moreover, even after vaccination plus two Omicron infections, the participant’s ACE2 competition and virus neutralization responses against BA.1 (as well as ACE2 competition activity against BA.2) plateaued at levels substantially lower than those seen against the wild-type strain, suggesting that the participant will remain at risk of new Omicron infections. A limitation of our study is that it did not assess T cell responses, which can reduce disease severity but may have less impact on virus transmission (53, 54), and thus we may be underestimating the protection that results from infection and reinfection in this case. Indeed, the participant’s symptoms following both infections were not severe enough to require hospitalization, demonstrating that vaccination was effective in its primary goal of preventing disease. Our results nevertheless illustrate the potentially limited ability of current vaccines to prevent recurrent infections and symptomatic disease caused by Omicron variants.

## Data Availability

Sequences were submitted to GISAID. The raw data supporting the conclusions of this article will be made available by the authors upon reasonable request.

## AUTHOR CONTRIBUTIONS

HRL and FM contributed equally as first authors. MGR, MAB and ZLB obtained project funding and contributed equally as senior authors. HRL, FM, MAB and ZLB designed the study. HRL, FM, PKC, YS, FY, RK, SD, RW, GU, SE, LY, WD, DK, and LB contributed to specimen collection and data analysis. HRL, YS, VL, DH, MLD, JS, NM, JSGM, CJB, NP, MN, CFL, MGR, MAB, and ZLB supervised the research and contributed to project management. HRL, MAB and ZLB wrote the original manuscript draft. All authors reviewed and approved the submitted version of the manuscript.

## ACKNOWLEDGEMENTS

We thank the leadership and staff of Providence Health Care for their support of this study. We thank the phlebotomists and laboratory staff at St. Paul’s Hospital, the BC Centre for Excellence in HIV/AIDS, the Hope to Health Research and Innovation Centre, and Simon Fraser University for assistance. Above all, we thank the participants, without whom this study would not have been possible.

## CONFLICT OF INTEREST

The authors declare that the research was conducted in the absence of any commercial or financial relationships that could be construed as a potential conflict of interest.

**Figure S1.**
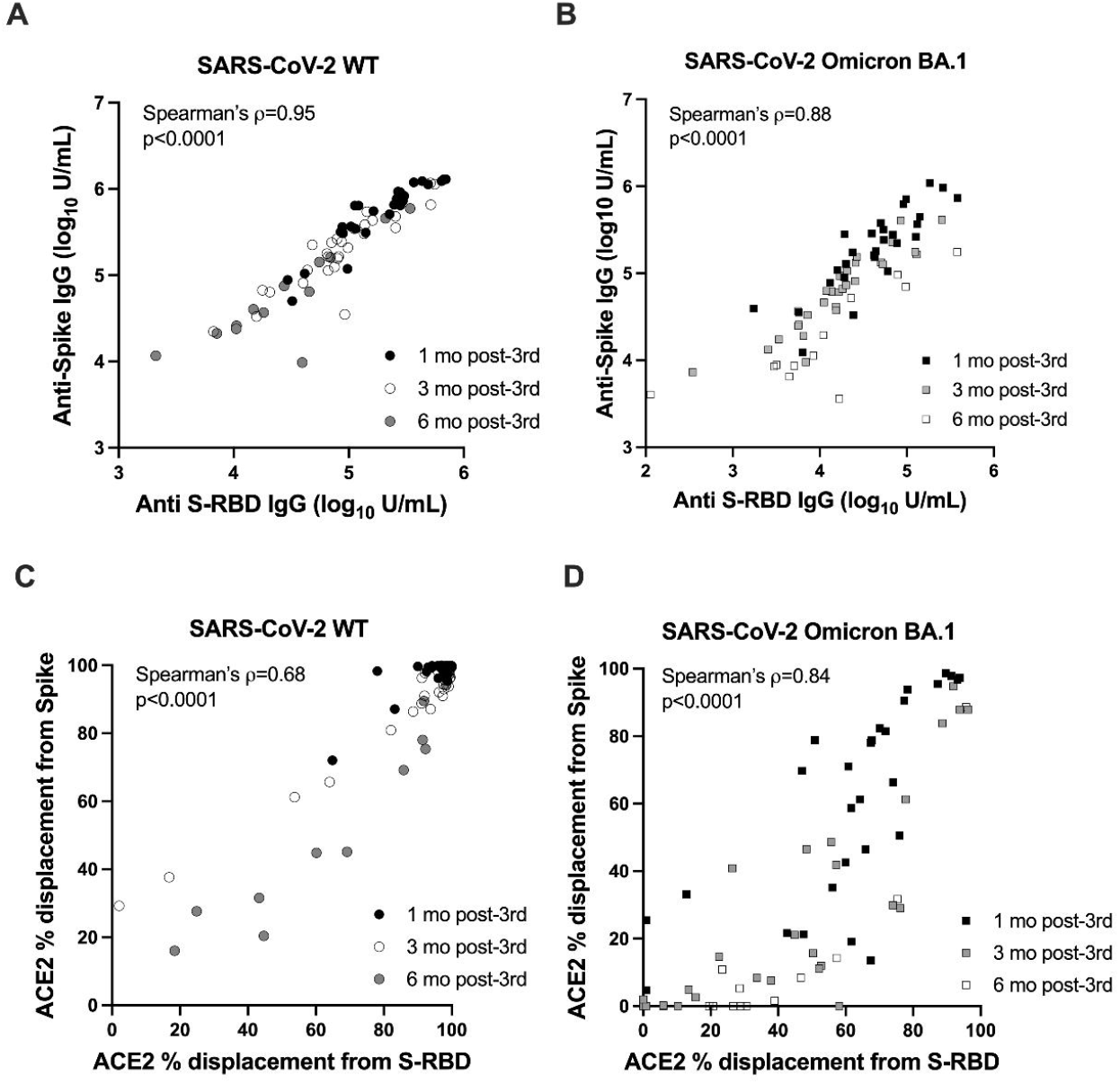
Correlations between wild-type (WT) and BA.1-specific anti-S-RBD and anti-Spike humoral responses measured following three COVID-19 vaccine doses using Meso Scale Diagnostics V-plex panels 22 and 25. All participants who were evaluated for BA.2 responses (i.e. those shown in Figure 2) are included in this analysis. Symbols are coloured based on post-vaccination time point, though the Spearman’s rho (ρ) and p-value reported are for the combined data.

